# In-home validation of wrist- and waist-worn devices against portable electroencephalography for sleep assessment in older adults

**DOI:** 10.1101/2025.10.28.25338962

**Authors:** Naoki Deguchi, Sho Hatanaka, Kaori Daimaru, Tomoko Wakui, Satoko Fujihara, Keigo Imamura, Hisashi Kawai, Kazushi Maruo, Hiroyuki Sasai

**Affiliations:** Research Team for Promoting Independence and Mental Health, Tokyo Metropolitan Institute for Geriatrics and Gerontology, 35-2 Sakae-cho, Itabashi-ku, Tokyo 173-0015, Japan; Research Team for Human Care, Tokyo Metropolitan Institute for Geriatrics and Gerontology, 35-2 Sakae-cho, Itabashi-ku, Tokyo 173-0015, Japan; Department of Biostatistics, Institute of Medicine, University of Tsukuba, 1-1-1 Tennodai, Tsukuba, Ibaraki 305-8575, Japan

**Keywords:** actigraphy, sleep assessment, older adults, portable electroencephalography, sleep parameters, total sleep time, sleep efficiency

## Abstract

Sleep health is essential for older adults. However, validity of wrist- and waist-worn devices for assessing sleep under free-living conditions remains unclear. This study evaluated the accuracy of a wrist-worn smartwatch (Silmee W22) and a waist-worn activity monitor (MTN-221) in measuring key sleep parameters, using portable electroencephalography (EEG; Insomnograf K2) as the reference. Healthy older adults wore all devices simultaneously for at least three nights. Total sleep time, sleep onset latency, wake after sleep onset, and sleep efficiency were analyzed using Bland– Altman plots, multilevel models, and intraclass correlation coefficients (ICCs). Fifty-five participants completed the study, yielding valid EEG-paired data for 49 participants with Silmee W22 (238 nights) and 53 with MTN-221 (265 nights). Silmee W22 overestimated total sleep time by 35 min and sleep efficiency by 8.1%, whereas MTN-221 overestimated it by 3 min and sleep efficiency by 1.0%. Both devices underestimated sleep onset latency and wake after sleep onset, with greater discrepancies observed as the estimated values increased. ICCs for total sleep time were 0.60–0.75 for Silmee W22 and 0.66–0.79 for MTN-221, while agreement for sleep onset latency and wake after sleep onset remained lower. While Silmee W22 did not provide sufficiently accurate estimates of total sleep time, MTN-221 yielded estimates that may offer practical benefits for large-scale sleep monitoring in older adults. In both devices, estimates of sleep onset latency, wake after sleep onset, and sleep efficiency should be interpreted with caution due to misclassification of quiet wakefulness. Further algorithm refinement is warranted.

## 1. Introduction

Sleep is fundamental to numerous physiological processes, including cognitive function, metabolism, appetite regulation, immune response, hormonal balance, and cardiovascular health. Aging is associated with characteristic changes in sleep architecture, such as a reduction in slow-wave sleep, increased nighttime awakenings, and earlier sleep onset, all of which contribute to a higher prevalence of sleep disorders, including insomnia and sleep apnea, among older adults (Li, Vitiello, & Gooneratne, 2022). Furthermore, sleep disturbances and difficulties initiating sleep have been linked to adverse health outcomes and a reduced quality of life in this population (Rodriguez, Dzierzewski, & Alessi, 2015). Traditional sleep assessment in older adults has often relied on self-reported measures. However, discrepancies between subjective self-reported sleep assessments and objective sleep measurements have been widely reported (Masaki et al., 2025). For instance, healthy sleepers tend to overestimate their sleep duration when using subjective assessments rather than objective measurements (Benz et al., 2023). This discrepancy suggests that potential sleep deprivation may go unnoticed. This highlights the need for accurate and objective sleep assessment methods to better support the health of older adults.

Polysomnography is the gold standard for sleep assessment but is often impractical for routine or long-term monitoring due to its reliance on specialized equipment and trained personnel (Jafari & Mohsenin, 2010; Newell, Mairesse, Verbanck, & Neu, 2012). As an alternative, portable electroencephalography (EEG) has gained recognition for enabling multi-night recordings in home environments, while accounting for individual differences in lifestyle factors, such as work schedules, physical activity, and habitual sleep patterns. Portable EEG achieves sleep stage classification accuracy comparable to polysomnography across various populations (de Gans et al., 2024; De Vos, Gandras, & Debener, 2014; Debener, Minow, Emkes, Gandras, & de Vos, 2012). Given its convenience and accessibility, portable EEG allows for extended monitoring under naturalistic conditions, facilitating more representative assessments of sleep in participant’s home environment.

However, although portable EEG is suitable for research purposes, it may still present usability challenges for widespread adoption in the general older adult population.

Growing societal interest in sleep monitoring has contributed to the widespread use of wearable devices equipped with accelerometers and heart rate sensors (Sadek, Demarasse, & Mokhtari, 2020). These devices estimate key sleep parameters—such as total sleep time (TST), sleep onset latency (SOL), wake after sleep onset (WASO), and sleep efficiency—at a low cost and with minimal user burden, making them well-suited for long-term sleep tracking (Chinoy et al., 2021). Their ease of use and ability to continuously collect both daytime activity and nocturnal sleep data make them particularly attractive for use among community-dwelling older adults. Consequently, wrist-worn devices and waist-worn devices are increasingly being utilized in research and health management (Kononova et al., 2019; Master et al., 2022; Straiton et al., 2018). Despite their growing popularity, the accuracy of these wearable devices in evaluating sleep characteristics—such as TST, WASO, SOL, and sleep efficiency—has not been sufficiently validated in older adults (Migueles et al., 2017). To fully leverage their potential in clinical and public health contexts, it is essential to establish their validity against objective reference measures such as portable EEG. Further research is needed to confirm the validity of sleep metrics obtained from wearable devices in this population (Baumert et al., 2022).

Therefore, this study aimed to evaluate the validity of wrist- and waist-worn accelerometer-based devices in measuring key sleep parameters in older adults, by comparing their outputs with those from a validated portable EEG device. The findings may inform the development of scalable and objective sleep assessment methods for both clinical and epidemiological research.

## 2. Methods

### 2.1 Design and settings

Data were collected as part of an ancillary study within the Smart Watch Innovation for Next Geriatrics & Gerontology (SWING-Japan) study (Shimura et al., 2025). SWING-Japan aims to promote health in older adults by utilizing the physical activity and sleep data obtained from wearable devices.

This ancillary sleep validation study was conducted from May to June 2023 and recruited participants from the Itabashi Longitudinal Study on Aging (Itabashi LSA), one of the cohorts included in the SWING-Japan project. The Itabashi LSA, conducted in February 2023 at the Tokyo Metropolitan Institute for Geriatrics and Gerontology, is an ongoing cohort study of community-dwelling older adults aged 70–85 years living in Itabashi Ward, located in the northwest area of Tokyo’s 23 special wards (Hatanaka et al., 2024).

### 2.2 Participants

Eligible participants were community-dwelling adults aged 70–85 years who had participated in the Itabashi LSA. Individuals were excluded if they met any of the following conditions: (1) a clinical diagnosis of insomnia according to the International Classification of Sleep Disorders, (2) suspected sleep disorders indicated by Pittsburgh Sleep Quality Index scores of >6 (Doi et al., 2000), (3) an average nightly continuous sleep duration of less than 6 h or more than 9 h, (4) a body mass index below 18.5 or equal to or above 30.0 kg/m², (5) a diagnosis of dementia or cognitive impairment (Mini-Mental State Examination score <23) (Shimura et al., 2025), (6) a diagnosis of depression or depressive symptoms (Geriatric Depression Scale-15 score >10) (Hatanaka et al., 2024), or (7) current smoking.

Potential participants were identified using data from the Itabashi LSA. They were mailed a brief study overview and a short screening questionnaire to assess additional eligibility criteria and willingness to participate. The questionnaire included questions regarding the following: (1) reported difficulty wearing wrist- and waist-worn devices and portable EEG device for five consecutive nights or recording their sleep schedules; (2) had used any type of sleep medications (prescription or over-the-counter) within the past month; (3) were unable to refrain from caffeine intake within five h before bedtime or from alcohol consumption during the study period; (4) shared a sleeping area with other individuals or animals; (5) had plans to stay overnight outside their home during the study period; or (6) had worked night shifts within the past month or were scheduled to do so during the study. Those who returned the questionnaire and met all eligibility requirements were invited to an orientation session, where they received further explanation and provided written informed consent.

At the end of the data collection period, participants received a 10,000-yen (approximately 67 dollars at the current exchange rate) gift card as compensation for their participation. This compensation was provided regardless of the amount or quality of data collected, to ensure that participation remained voluntary.

All procedures complied with relevant laws and institutional guidelines and were approved by the Research Ethics Committee of Tokyo Metropolitan Institute for Geriatrics and Gerontology on June 5, 2024 (Approval No.: R022-099). The study adhered to the Declaration of Helsinki. Written informed consent was obtained from all participants, and their privacy rights were fully protected.

### 2.3 Participant Characteristics measurement

Demographic and clinical assessment data were collected during the Itabashi LSA. Participants self-reported their age, sex, living arrangement, employment status, subjective health, Pittsburgh Sleep Quality Index (Doi et al., 2000), and 15-item Geriatric Depression Scale (Sugishita, Sugishita, Hemmi, Asada, & Tanigawa, 2017). Mini-Mental State Examination scores were assessed by trained personnel (Sugishita, 2010).

### 2.4 Portable EEG Device (reference)

Sleep variables were recorded using the portable EEG device Insomnograf K2 (S’UIMIN Inc., Tokyo, Japan) (**Figure 1A**). This lightweight device (162 g) employs flexible adhesive electrodes that are easy to attach and remove, making it particularly suitable for older adults. The device’s concordance with conventional polysomnography has been validated, with a κ coefficient of 0.71 for the classification of five sleep stages: Wake, rapid eye movement sleep (REM), stage 1 of non-REM sleep (NREM1), stage 2 of non-REM sleep (NREM2), and stage 3 of non-REM sleep (NREM3) (Seol et al., 2024).

**Figure 1.**
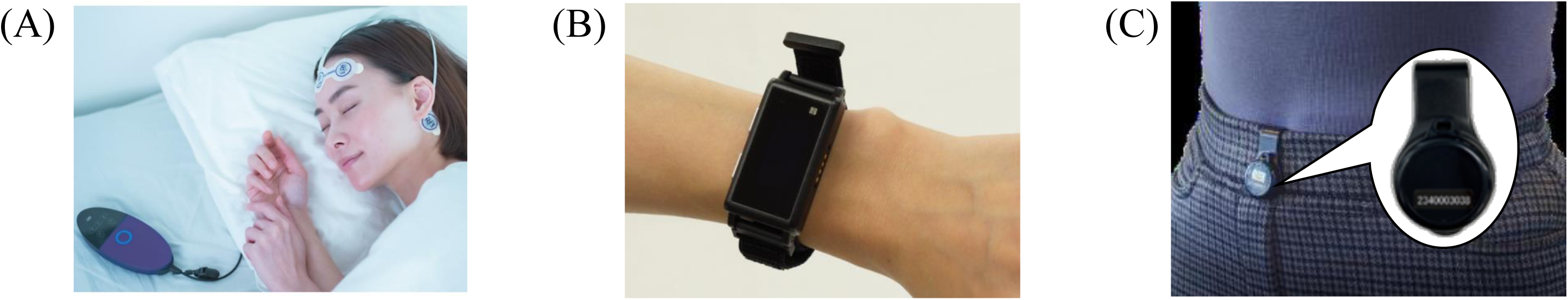
Images of (A) InSomnograf K2®* (S’UIMIN Inc., Japan), (B) Silmee W22 (TDK Corporation, Japan), **and (C) MTN-221 (ACOS Corporation, Japan).** *Publicly available at https://www.suimin.co.jp/

The recording system follows the international 10–20 system and includes four EEG electrodes (Fp1, Fp2, M1, and M2) and one reference electrode (Fpz). Sleep-stage analysis utilized four EEG derivations (Fp1–M2, Fp2–M1, Fp1–Mean M, Fp2–Mean M), with Fp1–Fp2 and Fp2–Fp1 serving as left and right electrooculograms, respectively, and M1–M2 as the submental electromyogram.

### 2.5 Wrist- and waist-worn device

#### 2.5.1 Wrist-worn device

The Silmee W22 (TDK Corporation, Japan) is a wrist-worn IoT device developed primarily for clinical and healthcare research in Japan (Yamada et al., 2025) (**Figure 1B**). Unlike consumer-grade smartwatches, it has been distributed mainly through business-to-business channels. The device is equipped with multiple sensors, including a triaxial accelerometer, photoplethysmography sensor, and skin temperature sensor. It also includes onboard memory and Bluetooth connectivity. These components enable continuous recording of raw data on physical activity, sleep–wake patterns (with sleep detection based on accelerometer-derived body movements), and physiological parameters such as heart rate and skin temperature. Data can be exported in CSV format for subsequent analysis. This compact and lightweight (52 × 24.5 × 14.0 mm; 26 g including the band) device can store approximately 30 days’ worth of data and is intended for continuous wear, except during charging or bathing. Although large-scale validation studies are limited, the Silmee W22 was selected for this study due to its strong technical support in Japan and its ability to provide raw sensor data—key requirements for high-quality data collection in research contexts. Although the Silmee W22 is no longer in production, its core technologies and sleep analysis algorithms have been inherited by the MiMoRy device (FrontAct Co., Ltd., Japan).

#### 2.5.2 Waist-worn accelerometer

The MTN-221 (ACOS Corporation, Japan) is a waist-mounted activity monitor designed specifically for institutional and research applications (Nakazaki et al., 2014) (**Figure 1C**). This small and lightweight device (diameter: 27 mm; thickness: 9.1 mm; weight: 9 g) contains a triaxial accelerometer that records data on physical activity and posture in 2-min intervals. It also detects body posture (e.g., standing, supine, prone, lateral) based on device orientation, enabling more accurate differentiation between activity and rest states. The MTN-221 has been evaluated in some validation studies involving younger adults and children (Nakazaki et al., 2014).In younger adults, the device was tested against polysomnography (PSG), with agreement, sensitivity, and specificity calculated for each sleep stage (wake, REM, stages 1, 2, and 3+4). These studies reported ∼85% overall agreement with PSG and comparable sensitivity and specificity across groups, confirming accuracy under controlled conditions. However, validation in older populations remains limited. The MTN-221 is particularly suitable for older adults because it is lightweight, unobtrusive, and equipped with a long-lasting battery that does not require charging during extended use, thereby minimizing participant burden and data loss. Therefore, the present study extends validation of the MTN-221 to an older adult population in community settings.

### 2.6 Data collection procedure

Based on prior studies suggesting that more than five nights of measurement are required for reliability and reduced errors (Sadeh, 2011), participants were requested to wear the devices for five consecutive nights whenever possible, and those with at least three valid nights of data were included in the analysis. Participants were instructed to wear all devices—portable EEG, Silmee W22, and MTN-221— each night before going to bed and to collect sleep data for a minimum of 5 days and up to 7 days. During this period, participants recorded "light-off time" and "wake-up time" in designated sleep diaries to calculate the time in bed (TIB).

The portable EEG was to be worn before bedtime. Participants were instructed to attach the electrodes after bathing or showering, press the record button before sleep to start the recording, and press the stop button upon waking. The Silmee W22 was worn on the dominant wrist at all days, except during water-related activities (such as showering, bathing, or swimming) or contact sports. The MTN-221 was clipped to the front of their torso after participants completed their bedtime routine. The participants were instructed to complete their bedtime routine at least 30 min before going to bed to improve the device’s ability to distinguish between sleep and wakeful states.

All devices were distributed after participants received instructions on their use at the time of enrollment. Participants returned all devices to the research team after completing the data collection period.

### 2.7 Data Processing

Data were synchronized across the portable EEG, Silmee W22, and MTN-221 based on their respective epoch-level classifications, using the first 60-s epoch. For all devices, scoring was conducted independently and blinded to other results. An independent researcher subsequently imported the epoch-level data into R (version 4.2.2) and aggregated them to calculate the following standardized sleep indices: (a) TST: total minutes scored as "sleep" within TIB; (b) SOL: time from lights-off to sleep onset; (c) WASO: time scored as "wake" from sleep onset to final awakening; and (d) sleep efficiency (%) = TST / TIB×100. SOL was defined as the time from "lights-off" to the first epoch classified as "sleep," where "sleep" was operationalized as the absence of any "wake" epochs for at least one consecutive minute (Dunican et al., 2018; Markwald, Bessman, Reini, & Drummond, 2016).

For the portable EEG data, a sleep EEG analyst with over 5 years of experience scored each 60-s epoch into five states: wake, REM, NREM1, NREM2, and NREM3. Epochs classified as REM or any non-REM stage were aggregated and treated as “sleep,” while Wake epochs were treated as “wake.” Scoring was conducted blinded to other results, and the datasets were subsequently linked by an independent researcher. For the Silmee W22, the collected data were analyzed using Silmee Pro Wx software (version 2.1.0.0; TDK Corporation, Japan) with the manufacturer’s proprietary algorithm, which classifies each 60-s epoch as either “wake” or “sleep” based on accelerometer-derived activity patterns. The epoch-level data are classified into binary states (wake/sleep), and some computational details, such as the internal algorithms and device operating frequencies, are not publicly disclosed.

Sleep indices were derived from these epoch-level classifications using the same definitions as those applied to the portable EEG data.

For the MTN-221, data were analyzed using Sleep Sign Act ver. 2.0 (Kissei Comtec Co., Ltd., Japan). Sleep and wake states were classified in 2-min epochs based on the default settings of a previously validated algorithm (Nakazaki et al., 2014). Similar to the Silmee W22, the output consists of binary epoch-level classifications (wake/sleep).

### 2.8 Statistical Analysis

Continuous variables were summarized as mean ± standard deviation, while categorical variables were reported as frequencies (percentages). In this study, sleep parameters obtained from the portable EEG and each wearable devices—Silmee W22 (wrist-worn) and MTN-221 (waist-worn)— including total TST, SOL, WASO, and sleep efficiency, were analyzed separately and evaluated for validity by comparison with the EEG reference.

An intercept-only multilevel model was used to calculate the overall mean and standard error for each sleep parameter, accounting for the hierarchical data structure (up to six valid nights per participant). Missing values exceeding 5% during a single night were addressed by excluding the data for those nights from the analysis. Only participants with data for at least five nights were included in the analysis (Bland & Altman, 2007).

To evaluate the agreement between the wearable devices and the reference EEG measurements, we calculated intraclass correlation coefficients (ICCs) and constructed Bland–Altman plots (Ludbrook, 2002). ICCs quantified the overall consistency of the measurements, while Bland–Altman plots provided a detailed view of systematic bias and the range of individual-level discrepancies. ICCs were calculated using a two-way mixed-effects model under the assumption of absolute agreement, for both single and average measures. These ICCs were used to estimate the agreement of the sleep parameters obtained from the wearable devices (Silmee W22 and MTN-221) relative to the portable EEG. The ICCs were interpreted as follows: “excellent” (≥0.75), “fair to good” (0.40–0.75), and “poor” (<0.40) (Rosner, 2006). Separate Bland–Altman analyses were conducted for Silmee W22 and MTN-221. In addition, intercept-only multilevel models were applied to estimate the overall mean of the paired differences between the portable EEG and each wearable device, along with the 95% limits of agreement (LoAs). Proportional bias was examined by including the mean of the two devices as a fixed effect and evaluating the slope coefficient. For non-normally distributed variables, a logarithmic transformation was applied before assessing the proportional bias.

Statistical analyses were conducted using SPSS 29.0 (IBM Corp., USA) and R version 4.2.2.

Statistical significance was set at a two-sided P value of < 0.05.

## 3. Results

Of the 654 participants in the Itabashi LSA, 479 were excluded, and invitations were sent to the remaining 175 individuals. Among these, 72 expressed willingness to participate, and 55 individuals were ultimately included in the study (**Figure 2**). The final analysis included 238 nights of data from 49 participants who used both EEG and the Silmee W22, as well as 265 nights of data from 53 participants who used both EEG and the MTN-221.

**Figure 2.**
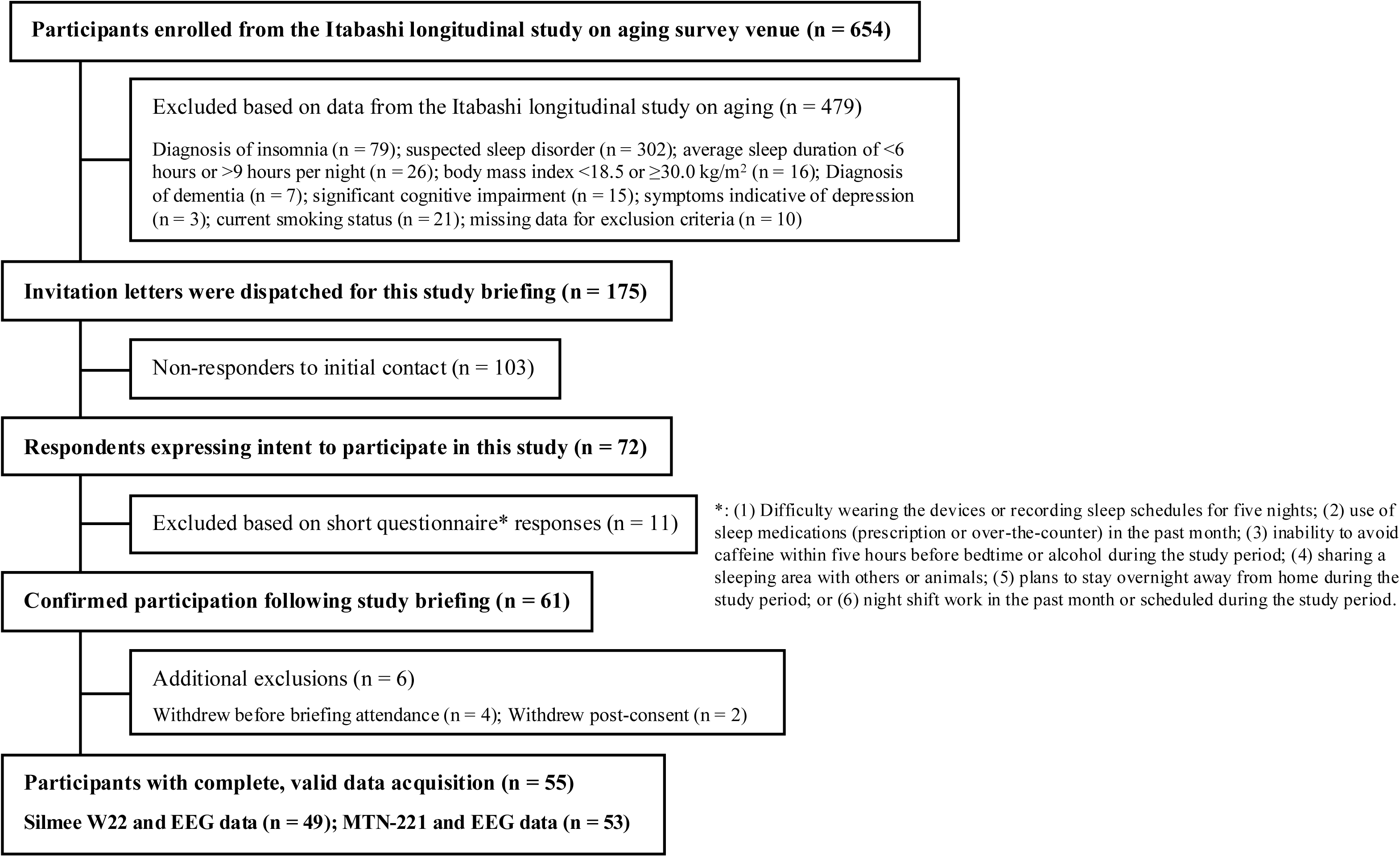
**Study Participant Flow**

The study sample was predominantly male with a mean age in the mid-70s and an average BMI of approximately 23. Approximately 20% lived alone, and educational attainment was evenly distributed between secondary and higher education levels. Most participants were employed in part-time or unstable jobs (**Table 1**).

**Table 1.**
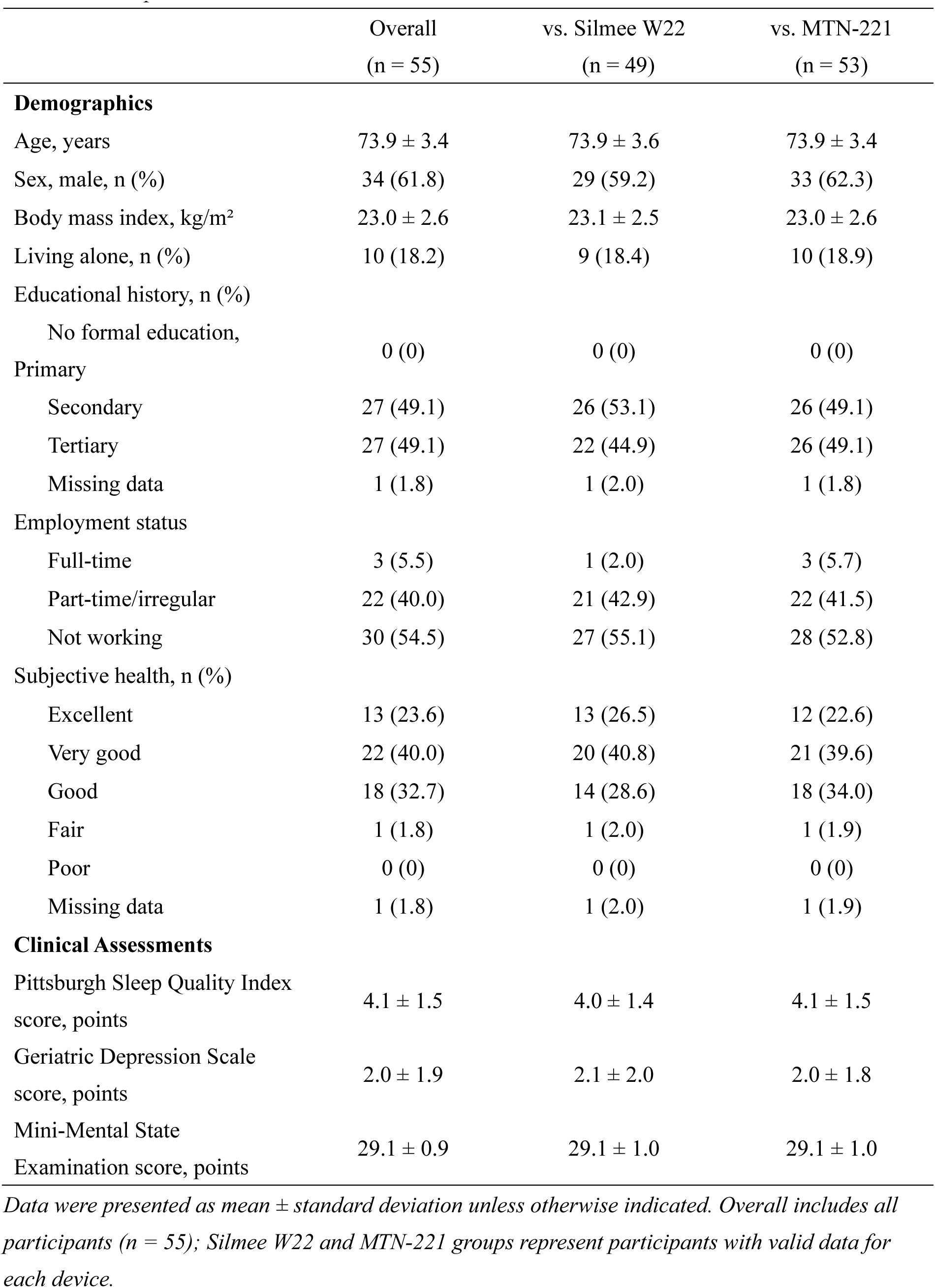
Participant characteristics

Among the 49 participants included in the comparison with the Silmee W22, the average TST, SOL, WASO, and sleep efficiency from the portable EEG were 336.1 min, 13.8 min, 58.1 min, and 80.2%, respectively. Among the 53 participants included in the comparison with MTN-221, the corresponding values were 340.6 min for TST, 13.7 min for SOL, 55.7 min for WASO, and 80.7% for sleep efficiency (**Table 2**).

**Table 2.**
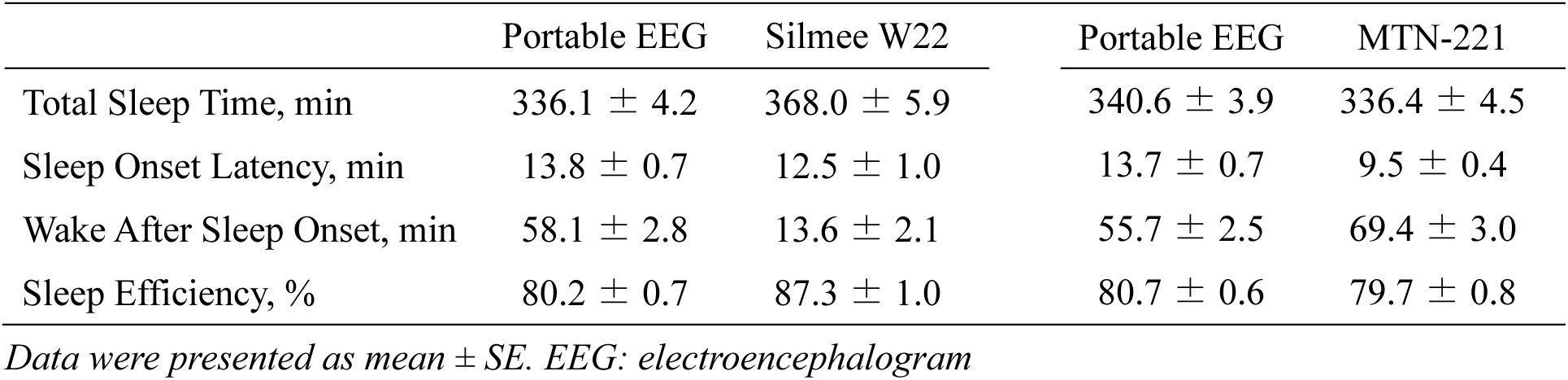
Descriptive sleep parameters for portable EEG, Silmee W22, and MTN-221

**Table 3.** Intraclass correlation coefficients between wearable devices and portable EEG across

| Sleep parameters | Single ICC (95% CI) | Average ICC (95% CI) |
| --- | --- | --- |
| vs. Silmee W22 |  |  |
| Total Sleep Time | 0.60 (0.44, 0.71) | 0.75 (0.61, 0.83) |
| Sleep Onset Latency | 0.14 (0.01, 0.26) | 0.24 (0.02, 0.41) |
| Wake After Sleep Onset | 0.31 (-0.03, 0.55) | 0.47 (-0.06, 0.71) |
| Sleep Efficiency | 0.32 (0.17, 0.45) | 0.49 (0.30, 0.62) |
| vs. MTN-221 |  |  |
| Total Sleep Time | 0.66 (0.58, 0.72) | 0.79 (0.74, 0.84) |
| Sleep Onset Latency | 0.18 (0.07, 0.30) | 0.31 (0.12, 0.46) |
| Wake After Sleep Onset | 0.36 (0.25, 0.46) | 0.53 (0.40, 0.63) |
| Sleep Efficiency | 0.27 (0.15, 0.38) | 0.42 (0.27, 0.55) |

*CI: confidence interval. ICC: intraclass correlation coefficients*
*Values are shown for single and average measurements.*

Bland–Altman analyses (**Figure 3**) showed that, compared with portable EEG, the Silmee W22 overestimated TST by an average of 35 min (LoA = -77 to 148). SOL was underestimated by 1.5 min (LoA = -40 to 37). WASO was shorter by 41 min on average (LoA = -125 to 43). Sleep efficiency was overestimated by 8.1% (LoA = -19 to 35). For TST, SOL, and WASO, the difference increased as the mean estimates increased, indicating proportional bias. Significant regression slopes were observed for TST ( *β* = 0.377, standard error [SE] = 0.051, p < 0.001), SOL ( *β* = 0.238, SE = 0.091, p < 0.010), WASO ( *β* = -0.190, SE = 0.077, p = 0.015), and sleep efficiency ( *β* = 0.571, SE = 0.079, p < 0.001).

**Figure 3.**
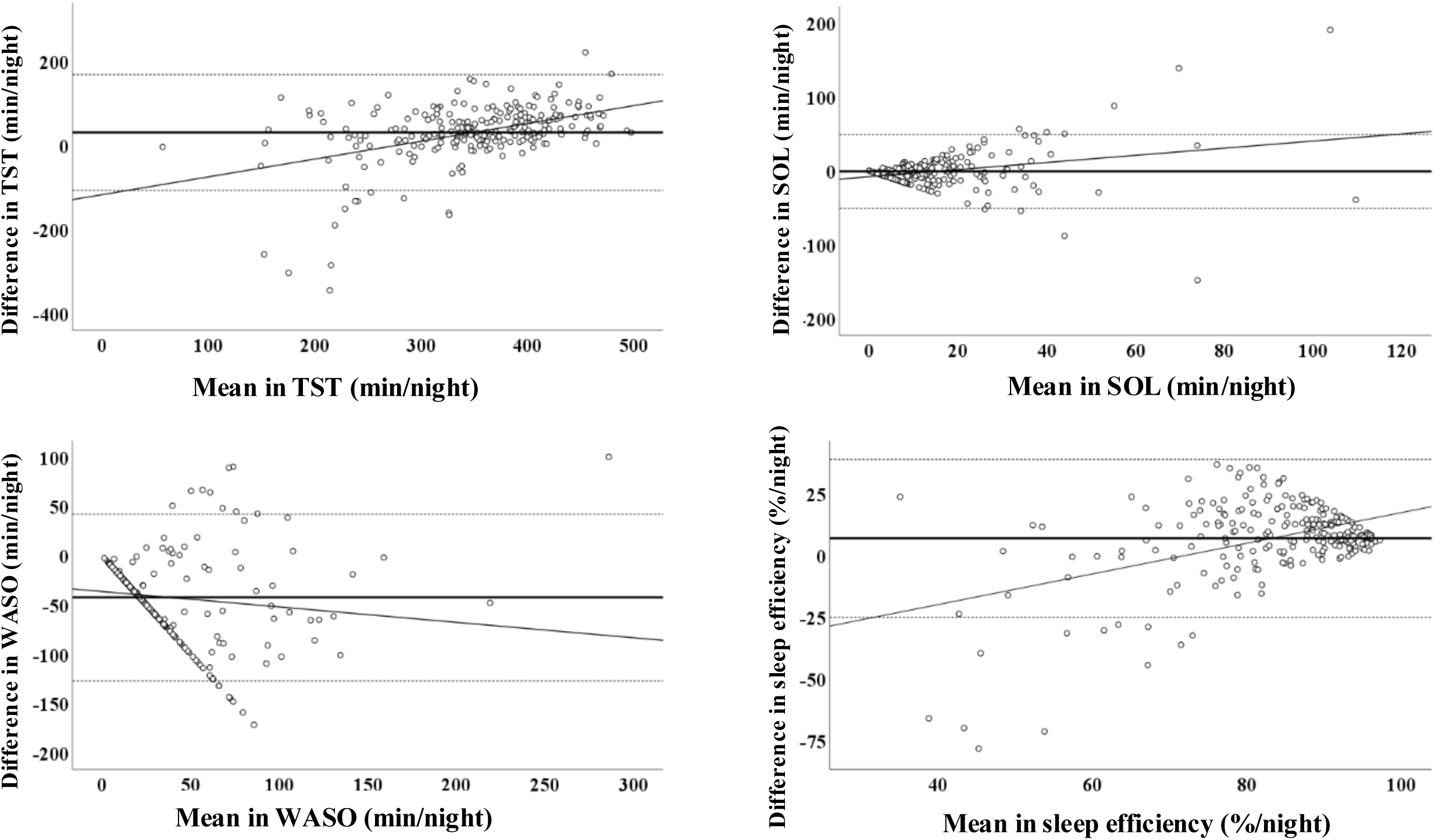
Bland–Altman plots comparing sleep parameters measured by the wrist-worn device (Silmee W22) and portable EEG. TST: Total Sleep Time, SOL: Sleep Onset Latency, WASO: Wake After Sleep Onset . The Bland–Altman plots compare sleep parameters derived from the wrist-worn device (Silmee W22) and portable EEG. The x-axis

In comparison with portable EEG devices, the MTN-221 device overestimated TST by 3 min (95% LoA: -118 to 112), underestimated SOL by 4.1 min (LoA: -27 to 19), underestimated WASO by 13 min (LoA: -88 to 114), and overestimated sleep efficiency by 1.0% (LoA: -29 to 27) (**Figure 4**). Similar to the Silmee W22, proportional bias was observed for MTN-221. Differences increased with the magnitude of the mean values, with significant regression slopes for TST ( *β* = 0.170, *p* = 0.003), SOL ( *β* = 1.066, *p* < 0.001), WASO ( *β* = 0.236, *p* = 0.004), and sleep efficiency ( *β* = 0.282, *p* = 0.003).

**Figure 4.**
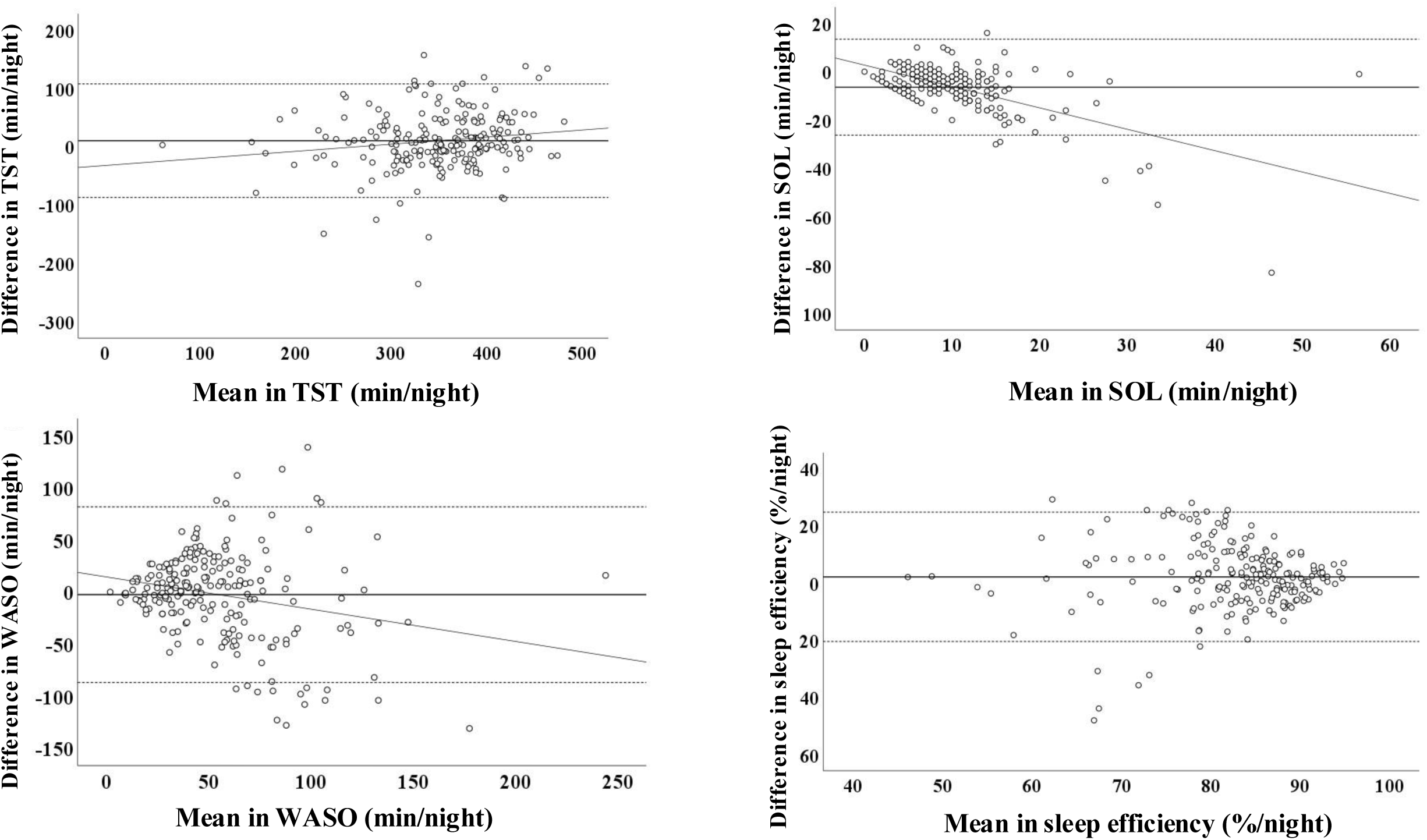
Bland–Altman plots comparing sleep parameters measured by the waist-worn device (MTN-221) and portable EEG. TST: total sleep time; SOL: sleep onset latency; WASO: wake after sleep onset. The Bland–Altman plots compare sleep parameters derived from the waist-worn device (MTN-221) and portable EEG. The x-axis

For single-night measurements, ICCs for TST were classified as “fair to good” for both Silmee W22 and MTN-221. In contrast, ICCs for SOL, WASO, and sleep efficiency were in the “poor” range. For average measurements, agreement improved. ICCs for TST reached the “excellent” range, while WASO and sleep efficiency achieved “fair to good” agreement. SOL remained in the “poor” category for both devices.

For the Silmee W22, the ICC for TST was 0.60 based on single-night data (“fair to good”) and improved to 0.75 with averaging (“excellent”). WASO showed moderate improvement from 0.31 to 0.47, and sleep efficiency from 0.32 to 0.49, both approaching the lower bound of “fair to good.” In contrast, SOL remained consistently low, indicating limited agreement across both measurement types.

For the MTN-221, the ICC for TST was 0.66 for single-night data (“fair to good”) and increased to 0.79 for averaged data (“excellent”). WASO rose from 0.45 to 0.56, within the “fair to good” range. Sleep efficiency improved from 0.27 to 0.42, reaching the threshold of “fair to good.” As with the Silmee W22, SOL remained low across both measurement types, reflecting poor agreement.

## 4. Discussion

In this study, we assessed the validity of two accelerometer-based wearable devices—Silmee W22 (wrist-worn) and MTN-221 (waist-worn)—for estimating sleep parameters in community-dwelling older adults. Validation was conducted in a real-world, in-home environment using portable EEG as the reference standard. Both devices showed moderate agreement with the portable EEG for TST, but marked discrepancies were observed for SOL, WASO, and sleep efficiency. Specifically, TST and sleep efficiency were consistently overestimated, whereas SOL and WASO were underestimated. Notably, the wrist-worn Silmee W22 exhibited larger deviations than those with the waist-worn MTN-221, suggesting that sensor placement and algorithm-specific sensitivity may contribute to systematic bias.

Both devices systematically underestimated WASO and SOL, aligning with previous findings that accelerometer-based devices often misclassify low-activity periods as sleep, thereby underestimating wakefulness (Plekhanova et al., 2023). Given the high prevalence of fragmented sleep and frequent nocturnal awakenings in older adults, optimizing the sensitivity of sleep–wake classification algorithms is essential. Bland–Altman analysis further indicated a proportional bias, whereby discrepancies between accelerometer-based devices and the portable EEG widened as sleep efficiency decreased. These findings highlight the importance of developing algorithms specifically tailored to the sleep characteristics of older adults, who typically experience shorter sleep durations and increased fragmentation (Full et al., 2018).

Consistent with previous findings using other wrist-worn devices (Waki et al., 2025), Bland– Altman analysis revealed a systematic bias in TST estimated from the Silmee W22. In the present study, the Silmee W22 consistently overestimated TST and sleep efficiency, while underestimating SOL and WASO. Errors increased in individuals with longer sleep duration and greater fragmentation. This pattern reflects a well-documented limitation of wrist actigraphy: high sensitivity for sleep detection but low specificity for wakefulness, leading to frequent misclassification of quiet wake as sleep (Chinoy et al., 2021; Guillodo et al., 2020). Guillodo et al. (2020) noted that such misclassification may be particularly pronounced in older adults, whose lower overall activity levels increases the likelihood of periods of quiet wakefulness being incorrectly scored as sleep. This age-related activity reduction may help explain the device’s reduced accuracies in classifying sleep–wake states. While TST estimates from the Silmee W22 were relatively stable when averaged over multiple nights, interpretation of SOL, WASO, and sleep efficiency at the individual level requires caution.

Future improvements may include integrating motion and cardiovascular signals, retraining algorithms using older adult data, and applying adaptive calibration techniques (Walch, Huang, Forger, & Goldstein, 2019).

The waist-worn MTN-221 demonstrated higher agreement and lower measurement bias across key sleep parameters, suggesting that waist-mounted devices—being less affected by extraneous limb movements—may provide more accurate sleep estimates (Inoue et al., 2019). Notably, multi-night assessments improved the validity of TST and WASO, yielding ICCs indicative of good agreement.

Given its small size, ease of use, and minimal participant burden, the MTN-221 shows a strong potential for large-scale epidemiological studies. However, single-night ICC values for WASO and sleep efficiency remained below established clinical thresholds (Tracy et al., 2014). These findings highlight the need for cautious interpretation in clinical contexts, particularly when evaluating SOL and WASO.

Our findings contribute to the growing body of evidence on both the capabilities and limitations of accelerometer-based wearable devices for sleep assessment in older adults. To improve accuracy in this population, future efforts should prioritize refinement of sleep detection algorithms and validation across a broader range of sleep patterns. Integrating additional physiological signals—such as heart rate variability and respiratory patterns—alongside accelerometer data may further enhance classification accuracy (Beattie et al., 2017). While wearable devices offer practical benefits for large-scale, in-home sleep monitoring, further calibration against EEG-based benchmarks remains essential to increase their precision for both clinical and epidemiological applications.

This study has several limitations. First, our sample consisted of older adults from a single geographic region, and the application of strict exclusion criteria, such as excluding current smokers and individuals using sleep medication, may limit the generalizability of our findings. These participants were excluded to minimize the influence of factors known to be associated with sleep disturbances, thereby ensuring that the validation focused on device performance under relatively stable sleep conditions. Second, as the devices’ internal algorithms were proprietary and not publicly available, the specific sources of measurement errors could not be ascertained. Third, although portable EEG is suitable for field-based sleep assessment, its diagnostic accuracy remains inferior to that of polysomnography. Given these limitations, future studies should employ larger and more diverse cohorts, longer monitoring durations, and direct comparisons with polysomnography to strengthen external validity.

Despite these constraints, the findings carry practical and clinical implications. The Silmee W22 may be suitable for tracking habitual sleep duration and long-term trends in older adults at the population level, but its use for evaluating night-to-night sleep continuity at the individual level should be approached with caution. In contrast, the MTN-221, with its better agreement and lower participant burden, appears more promising for large-scale epidemiological studies. In clinical contexts, wearable devices can serve as pre-screening tools or for longitudinal monitoring, but polysomnography remains essential for definitive diagnosis.

## 5. Conclusion

This study evaluated the accuracy of two accelerometer-based wearables—the wrist-worn Silmee W22 and the waist-worn MTN-221—for sleep assessment in older adults, using portable EEG as the reference standard. Both devices showed moderate agreement with portable EEG for TST, supporting their potential use in large-scale studies. However, both devices underestimated SOL and WASO, resulting in inflated estimates of sleep efficiency. These discrepancies likely reflect the misclassification of quiet wakefulness as sleep, a known limitation of accelerometry-based sleep monitoring tools. The waist-worn MTN-221 provided more accurate TST estimates, potentially due to reduced sensitivity to upper limb movements. Nonetheless, metrics of sleep continuity and fragmentation should be interpreted with caution, particularly in older adults who typically exhibit low movement during wakefulness. Future research should prioritize refining algorithms, extending monitoring durations, and validating these tools against polysomnography, especially when sleep continuity metrics are of clinical interest.

## Data Availability

All data produced in the present study are not available

## Glossary

EEG, electroencephalography; ICC, intraclass correlation coefficient; SOL, sleep onset latency; TIB, time in bed; TST, total sleep time; WASO, wake after sleep onset; Itabashi LSA, Itabashi Longitudinal Study on Aging; REM, rapid eye movement sleep; NREM1, stage 1 of non-REM sleep; NREM2, stage 2 of non-REM sleep; NREM3, stage 3 of non-REM sleep; SE, standard error; LoA, 95% limits of agreement

## Data Availability

The datasets generated and/or analyzed during the current study are available from the corresponding author upon reasonable request.

## Ethics approval statement

This study was approved by the Ethics Committee of Tokyo Metropolitan Institute for Geriatrics and Gerontology (approval number: R022-099).

## Patient consent statement

Written informed consent was obtained from all participants prior to their inclusion in the study.

## Permission to reproduce material from other sources

Permission to reproduce the figure from S’UIMIN Inc., TDK Corporation and ACOS Corporation has been obtained from the copyright holder.

## Acknowledgements

We would like to thank Editage for editing and reviewing this manuscript for English language.

## CRediT authorship contribution statement

Naoki Deguchi: Conceptualization, Formal analysis, Investigation, Methodology, Writing – original draft. Sho Hatanaka: Formal analysis, Writing – review & editing. Kaori Daimaru: Methodology, Project administration, Writing – review & editing. Tomoko Wakui, Satoko Fujihara, Keigo Imamura, and Hisashi Kawai: Prepared study devices, Participant recruitment, Writing – review & editing.

Kazushi Maruo: Supporting statistical analysis, Writing – review & editing. Hiroyuki Sasai: Conceptualization, Funding acquisition, Methodology, Supervision, Writing – review & editing.

